# A scoping review of virtual morning report and outcomes in Canada and the United States

**DOI:** 10.1101/2022.11.28.22282625

**Authors:** Shohinee Sarma, Tharsan Kanagalingam, James Lai, Tehmina Ahmad

**Affiliations:** Endocrinology and Metabolism, Mount Sinai Hospital, University of Toronto, Canada; Schulich School of Medicine, at Western University, Canada; Family Medicine, Orillia Soldiers’ Memorial Hospital, Canada; Endocrinology and Metabolism, Women’s College Hospital, University of Toronto, Canada

## Abstract

**Purpose:** To describe the current landscape of virtual morning report (VMR) in medical residency education including its varying formats, methods, and associated effectiveness on learning and clinical outcomes.

**Methods:** The authors conducted a scoping review using the Arksey and O’Malley methodology. They searched Embase, OvidMEDLINE, Google Scholar, and PubMed between January 1, 1991 to April 15, 2022. Articles written in English on virtual morning report and virtual case-based teaching in medical residency programs were captured. Two authors independently screened articles using the inclusion criteria. Using a snowball approach, further citations were identified from included references. Two authors performed data extraction including outcomes using the Kirkpatrick model. We conducted thematic analysis using an iterative process.

**Results:** A total of 401 citations were screened for eligibility and we included 40 articles. The number of published studies per year on VMR increased since the COVID-19 pandemic. Most studies used online case-based modules (n=20; 50.0%) or videoconferencing (n=12; 30.0%). The majority of studies described improved confidence with clinical reasoning, easy access, and preference for chatboxes/polls for engagement (Kirpatrick level 1). Nineteen studies demonstrated improved knowledge acquisition with pre-and post-test scores (Kirkpatrick level 2). Behaviour changes (Kirkpatrick level 3) included improved screening tests and medication prescribing. There were no studies on clinical outcomes (Kirkpatrick level 4). Thematic analyses revealed that VMR increased clinical reasoning, efficiently used technology, provided an inclusive environment for diverse learners, but reduced peer engagement and bedside teaching.

**Conclusion:** Virtual morning report has a positive impact on learner confidence, knowledge, inclusivity, accessibility, and behaviour change. Future research is needed to explore the impact on patient outcomes as well as identify strategies for peer engagement and social interaction.

## Introduction

The traditional morning report is a hallmark of medical education worldwide. It generally covers a wide range of topics including diagnostic dilemmas, rare cases, patient safety, ethics, evaluation of tests and procedures, and teaching around a core curriculum topic.^1,2,3,4^ Specifically, in internal medicine and medicine subspecialties, morning report has been a vital tradition that medical learners rank as one of their core educational activities in training with some implication on reduced hospital length of stay and hospitalization costs.^2,5,6^ The coronavirus-19 (COVID-19) pandemic has created a situational need to adapt to online forms of teaching and accelerated the utilization of virtual forms of morning report.^10^ Due to cancelled conferences during COVID-19, accessing a virtual network of colleagues is critical to promote opportunities for educational growth and career progression. On the other hand, patient privacy, blurring of workplace and home boundaries, and the challenges of identifying trustworthy sources are ongoing considerations in virtual teaching. We conducted a preliminary search of studies describing virtual morning report (VMR) and did not find a standard definition and interventions varied between studies.^11,12,13,^ Thus, the objective of this scoping review is to describe the current landscape of virtual morning report (VMR) and to identify its effectiveness on learning and clinical outcomes.

## Methods

We used the original scoping review method developed by Arksey and O’Malley and the Joanna Briggs Institute to conduct this scoping review.^14^ Our study included the following steps: 1) developing the research question and eligibility criteria; 2) searching, screening, and identifying relevant studies; 3) selecting studies to include; 4) charting data from included studies; and 5) collating data, identifying themes, and reporting results. We used the Preferred Reporting Items for Systematic Reviews and Meta-Analyses (PRISMA) Extension for Scoping Reviews guidelines

### Eligibility Criteria

#### Population

All learners in various stages of internal medicine, family medicine, and medicine subspecialty education within North America were included. These stages encompassed medical school, medicine residency programs, and medicine subspecialty fellowship programs.

#### Interventions

All interventions that included an online, audiovisual, or virtual format for morning report or case-based teaching or case-based simulation were included.

#### Outcomes

All relevant learning, teaching, and clinical outcomes were eligible for inclusion. Examples from preliminary literature review included learner perception, increased participation, communication, expanded knowledge base, patient safety, and clinical efficiency.^2,6,7^

#### Study designs

We included all peer-reviewed primary studies in the English language including randomized control trials (RCTs), observational studies, quasi-controlled studies, and qualitative reports.

#### Timeframe

Studies published after between January 1, 1991 to April 15, 2022 were eligible for inclusion to capture all eligible online programs since the world wide web was created. We excluded review articles without primary data, studies conducted prior to 1990, and studies conducted in surgical programs due to heterogeneity of teaching methods between medical and surgical specialties. We excluded studies that involved virtual instructional modules that were not case-based. We excluded systematic reviews, but they helped inform eligibility and search criteria.

### Information Sources and Strategy

We worked with an experienced library information specialist to develop the search strategy (Supplementary Appendix). We performed an initial search in two databases EMBASE and OvidMEDLINE using our search criteria (Supplementary Appendix) on August 30, 2021. We then searched for additional newer articles by referencing search engines Google Scholar and Pubmed with our search keywords (Supplementary Appendix) on April 15, 2022. We also used the snowball sampling approach to manually scan references from included studies and systematic reviews to find additional articles.

### Study Selection

We used the online Covidence tool (Covidence.org, Australia, version 2022) to import and collate citations for title, abstract, and full text screening outlined in the PRISMA flow diagram (Figure 1). Two independent reviewers (TK and SS) used the inclusion criteria to screen titles and abstracts (Level 1 screening) and full text articles (Level 2 screening). We completed a pilot exercise of five studies at each level of screen to ensure agreement between reviewers. Any discrepancies between reviewers were resolved by consensus after reading the full manuscript.

**Figure 1:**
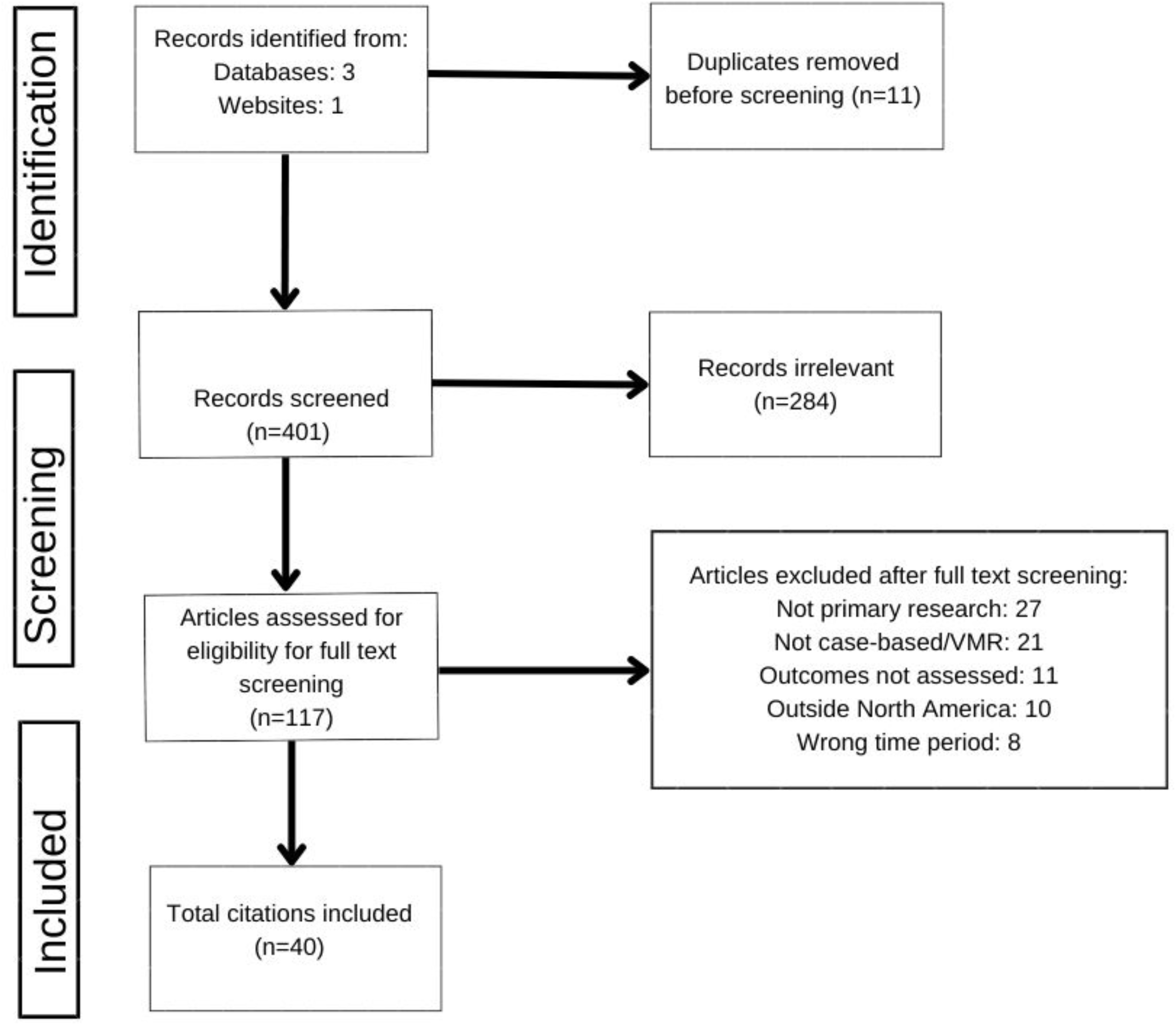
PRISMA flow diagram of included studies.

### Data Abstraction

We drafted a data abstraction form using Excel (Microsoft Excel 355, version 16.61, Microsoft Corporation, Redmond, Washington) including characteristics of the study (date of publication, authors, country of study, study aims, study population, and study design/interventions), learner characteristics (level of education, age, gender, sex), and outcomes (learning, teaching, and clinical outcomes). Two reviewers (TK and SS) piloted this form on five studies and then modified the form based on feedback to ensure clarity and agreement between reviewers. Two reviewers (TK and SS) independently extracted data from all studies meeting inclusion criteria after Level 2 screening. Any discrepancies were resolved by consensus. We did not conduct risk of bias assessments in keeping with the Joanna Briggs Institute Methods Manual for Scoping Reviews.^15^

### Data Synthesis

Our synthesis focused on describing the current landscape of virtual morning report and their effectiveness measures using Kirkpatrick’s four levels of evaluation.^16,17^ The educational and clinical effectiveness outcomes were categorized using Kirkpatrick’s levels of learning (reaction, learning, behaviour, and results) using Excel. We used an inductive^18^ and constructivist^19^ approach to thematic analysis for qualitative comments by learners. We extracted qualitative comments from studies and three authors (SS, TK, and TA) used a coding scheme on Dedoose (version 9.0.46, SocioCultural Research Consultants, Los Angeles, California) to create a coding tree. The coding tree was used to code all extracts by authors (SS, TK, and TA) and revised after discussion to ensure clarity, accuracy, and coding agreement. Emerging themes were defined after discussion between authors (SS, TK, and TA).

## Results

Our initial search of two databases EMBASE and OvidMEDLINE and additional searches of Google Scholar and PubMed search engines produced a total of 401 citations for title and abstract screening (Level 1 screen) (Figure 1). Of these, 117 citations were eligible for full text screening (Level 2 screening). We included a final 40 citations in our scoping review after Level 2 screening, which included 38 journal articles and 2 conference abstracts.^20,21^ Figure 1 outlines the PRISMA flow diagram of included studies after screening.

### Descriptive Characteristics

The number of studies on virtual morning report and case sharing per year increased significantly since 2020 after the global COVID-19 pandemic. One study (2.5%) was published in 2022,^20^ 7 studies (17.5%) were published in 2021,^2,22,23,24,25,26,27^ and 8 studies (20.0%) were published in 2020.^28,29,30,3,31,32,33,21^ The studies and their corresponding citations are available in Table 1.

**Table 1:** Descriptive characteristics of included studies.

| Characteristic | No. of articles (%) |
| --- | --- |
| **Countries** |  |
| United States of America | 34 (85.0%)<sup>2,20,21, 22, 23, 24, 25,26,28,29,30,31, 33,34,35,36,37,38,39,40,41,42,43,44, 45,46,47, 48,49, 50,51,52,53,60</sup> |
| Canada | 6 (15.0%)<sup>32,54,55,56,58,57</sup> |
| **Programs/specialties** |  |
| General internal medicine | 25 (62.5%)<sup>2,22,25,26,29,30,31,33,35,36,37,38,39,40,41,42,44,49,50,52,53,54,55,56,58</sup> |
| Family medicine | 4 (10.0%)<sup>21,37,39,40</sup> |
| Neurology | 4 (10.0%)<sup>32,57</sup> |
| Cardiology | 2 (5.00%)<sup>28,45</sup> |
| Nephrology | 1 (2.50%)<sup>42</sup> |
| Rheumatology | 1 (2.50%)<sup>56</sup> |
| Other programs | 2 (5.00%)<sup>34,39</sup> |
| **Level of training** |  |
| Medical students | 16 (40.0%)<sup>20, 21,23,25,26,30,31,34,37,43,46,47, 52, 54,56, 60</sup> |
| PGY1 resident | 14 (35.0%)<sup>2,24,25,35,41,43,48,51,53, 54,55,56,58,61</sup> |
| PGY2 resident | 16 (40.0%)<sup>2,24,25,29,35,36,38,41,48,51,53,54,55,56,58,61</sup> |
| PGY3 resident | 13 (32.5%)<sup>2,24,35,38,41,48,51,53, 54,55,56,58, 61</sup> |
| PGY4-5 resident | 7 (17.5%)<sup>21,41,48,51,54,61,62</sup> |
| Fellow | 4 (10.0%)<sup>23,26,62,61</sup> |
| **Publication year<sup>2</sup>** |  |
| 2000-2005 | 6 (15.0%)<sup>35,43,46,47,49,52</sup> |
| 2006-2010 | 7 (17.5%)<sup>36,37,38,39,51,58,60</sup> |
| 2011-2015 | 4 (10.0%)<sup>40,41,53,54</sup> |
| 2016-2020 | 15 (37.5%)<sup>21,28,29,30,31,32,34,42,44,45,48,55,56,57,61</sup> |
| 2021-2022 | 8 (20.0%)<sup>2,20,22,23,25,26,50,62</sup> |
| **Methodology** |  |
| Mixed method | 19 (47.5%)<sup>2,23,26,29,30,31,39,40,42,43,48,50,52,53,54,55,56,60,62</sup> |
| Randomized controlled trial | 10 (25.0%)<sup>34,35,36,37,38,41,46,47,51,58</sup> |
| Observational design | 3 (7.50%)<sup>21,44,49</sup> |
| Qualitative design | 2 (2.50%)<sup>22,25</sup> |
| Descriptive | 6 (15.0%)<sup>20,28,32,45,57,61</sup> |
| Disease topics |  |
| Diabetes management | 3 (7.50%)<sup>29,39,53</sup> |
| Osteoporosis | 1 (2.50%)<sup>41</sup> |
| Chronic pain | 1 (2.50%)<sup>44</sup> |
| Opioid prescribing | 1 (2.50%)<sup>51</sup> |
| Advanced care planning | 1 (2.50%)<sup>22</sup> |

Abbreviation: PGY=postgraduate year

#### Countries

The scoping review focused on programs within North America. The majority of included studies (34 studies) were conducted in the United States of America (85.0%)^2,28,34,29,22,30,35,36,37,38,39,40,40,41,42,43,23,31,44,45,24,46, 47,33,25,26,20,21,48,49,50,51,52,53^ and 6 studies^54,55,56,57,58,32^ were conducted in Canada (15.0%). Two studies had international impact with learners globally using the morning report blog and The Clinical Problem Solvers podcast (Table 1).^54,25^

#### Programs and specialties

Twenty-five studies (62.5%) were conducted among general internal medicine programs,^2,22,25,26,50,29,30,31,33,44,42,55,40, 41,54,53,39,37,36,58,35,49,52^ 4 studies (10.0%) included family medicine learners,^21,40,39,37,59^ 4 studies (10.0%) were conducted in neurology programs,^23,24,57,32^ 1 study in nephrology (2.5%),^42^ 1 study (2.5%) in rheumatology,^56^ 2 studies (5.0%) in cardiology,^28,45^ and 2 studies (5.0%) in other programs (Table 1).^34,48^

#### Level of training

Table 1 demonstrates the percentage of participants across medical school, residency, and fellowship.

#### Participant characteristics

Only six studies (15.0%) reported the number and/or proportion of participants by sex.^2,25,36,38,41,51^ There were no studies reporting on ethnicity or racial composition of learners.

#### Format for virtual morning report

Most studies (n=20; 50.0%) used an online computer-assisted instructional course or web modules to outline patient cases (Figure 2). ^29,34,35,36,37,38,39,40,41,43,44,46,47,48,49,51,52,53,56,60^ Twelve studies (30.0%) used a videoconferencing platform (e.g.: Zoom or Microsoft Teams) to share cases in real time with interaction. ^2,20,21,22,23,26,27,28,31,32,57,61^ The formats and proportion of programs are displayed illustratively in Figure 2.

#### Study aims and methodologies

Most studies (34 studies; 85.0%) focused on knowledge acquisition and learning effectiveness using a combination of qualitative, observational, randomized trials (RCT), and mixed method designs (Table 1). Table 1 provides further breakdown of study methodologies.

#### Study outcomes

Most studies explored the impact of virtual morning report (VMR) on knowledge acquisition and retention. ^21,23,25,29,30,34,35,36,38,39,40,41,42,43,44,46,47, 48,49,50,51,52,53,55,56,62,63,60,65^ Two studies also evaluated if VMR improved efficiency and time spent learning ^38,56^ and two studies explored association between the VMR format and cognitive learning styles.^36,37^ Six studies reported on clinical outcomes in the areas of inpatient diabetes management, osteoporosis, chronic back pain, opioid prescribing, and advanced care planning (Table 1).^22, 39,41,44,51,53^ The six descriptive studies did not evaluate any component of learning or clinical outcomes.^20,28,32,45,57,61^

### Learning and Clinical Outcomes (Kirkpatrick Levels)

#### Reaction

Overall, learners positively valued their experience with VMR with highlighted qualities of increased confidence,^21,23,26,44^, supportive learning community, and enhanced educational experience.^42,45,56^ Positive features also included increased accessibility from home, interaction online with peers and faculty with videoconferencing, and valuable patient discussions to bolster clinical reasoning and management.^25,41,53^ Harnessing the use of technology with chat boxes, audience response systems, and use of smartboard/tablets improved learner experience.^2,23,28,32,61^ There was also reduced fear of exposure to COVID-19 by lessening inpatient case teaching rounds.^62^

Increased diversity of learners in a safe online teaching environment was another positive feature.^2,25^ Learners preferred having easy access to the chat function to ask questions and post comments compared to increased fear of speaking aloud in traditional medical hierarchy-based environments.^25,28,32^ Many learners also felt that the virtual format positively supplemented traditional case-based teaching but could not replace bedside or clinical teaching.^44,62^ Finally, being able to return to video modules and podcasts later was valuable in reinforcing topics.^2^

Barriers and challenges to virtual environments included lack of engagement with lecture-based modules, reduced bedside teaching, lack of access to physical exams, and increased work preparing for online cases by learners and faculty.^29,44,62^ The switch to VMR also lessened feelings of social connectedness.^2^

#### Learning

Fifteen studies actively examined improvements in knowledge after VMR with pre-and-post test questions. ^23,29,34,36,38,39,40,41,43,48,49,51,52,53,63^ In most cases, mean post-test scores were higher than pre-test scores and knowledge retention occurred even with testing 6 months later.

For studies exploring two different formats for virtual case teaching (eg: adaptive vs. standard formats), there was no difference in test scores.^39,41,63^ In two studies exploring an association between VMR and cognitive styles, there was no difference in test scores or personal preference.^36,63^ There was no association between learning styles and use of adaptive multiple-choice questions or self-assessment questions to supplement virtual case-based teaching.^36^

#### Behaviour

Learners felt that VMR increased participation and attendance in case discussion compared to traditional formats.^60^ Some also reported heightened confidence in clinical scenarios with diagnosis and clinical reasoning, but these studies did not compare to traditional morning report. ^21,23,26,44^ Learners also reported feeling better organized and comfortable in conducting online oral presentations themselves after exposure to VMR discussions.^50^

For clinical behaviour change, learners that attended virtual case-based teaching on inpatient diabetes management were found to switch from use of sliding scale insulins to basal-bolus regimens and demonstrated improved comfort in managing hyperglycemia.^53^ Similarly, internal medicine residents who underwent modules on pain management and case-based palliative care teaching reported self-rated competence in the use of opioids for chronic pain management and increased comfort in advanced care planning discussions.^22^ There was also a higher percentage of documentation of physical examinations in clinical encounters for residents who viewed virtual case modules on chronic back pain.^44^ Residents who were randomized to case-based teaching on osteoporosis also calculated fracture risk assessments (FRAX) at a higher proportion compared to residents who did not receive this teaching.^41^

#### Results

Only one randomized study directly studied the impact of VMR on patient-related outcomes with increased screening bone densities and prescribing of bisphosphonates after osteoporosis case-teaching but did not directly assess change in fracture risk.^41^ None of the other studies reported structural change or patient outcomes.

### Thematic Analysis

We identified four central themes based on our thematic analysis of qualitative comments: 1) Improved clinical reasoning and knowledge; 2) Safety and inclusivity; 3) Accessibility and effective use of technology; and 4) Interaction and social wellbeing. We identified themes from analysis of 41 qualitative free-text excerpts from 21 studies that included a qualitative component (Table 2). ^2,22,25,26,27,29,30,34,35,38,39,42,46,47,49,50,53,56,57,60,62^

**Table 2:**
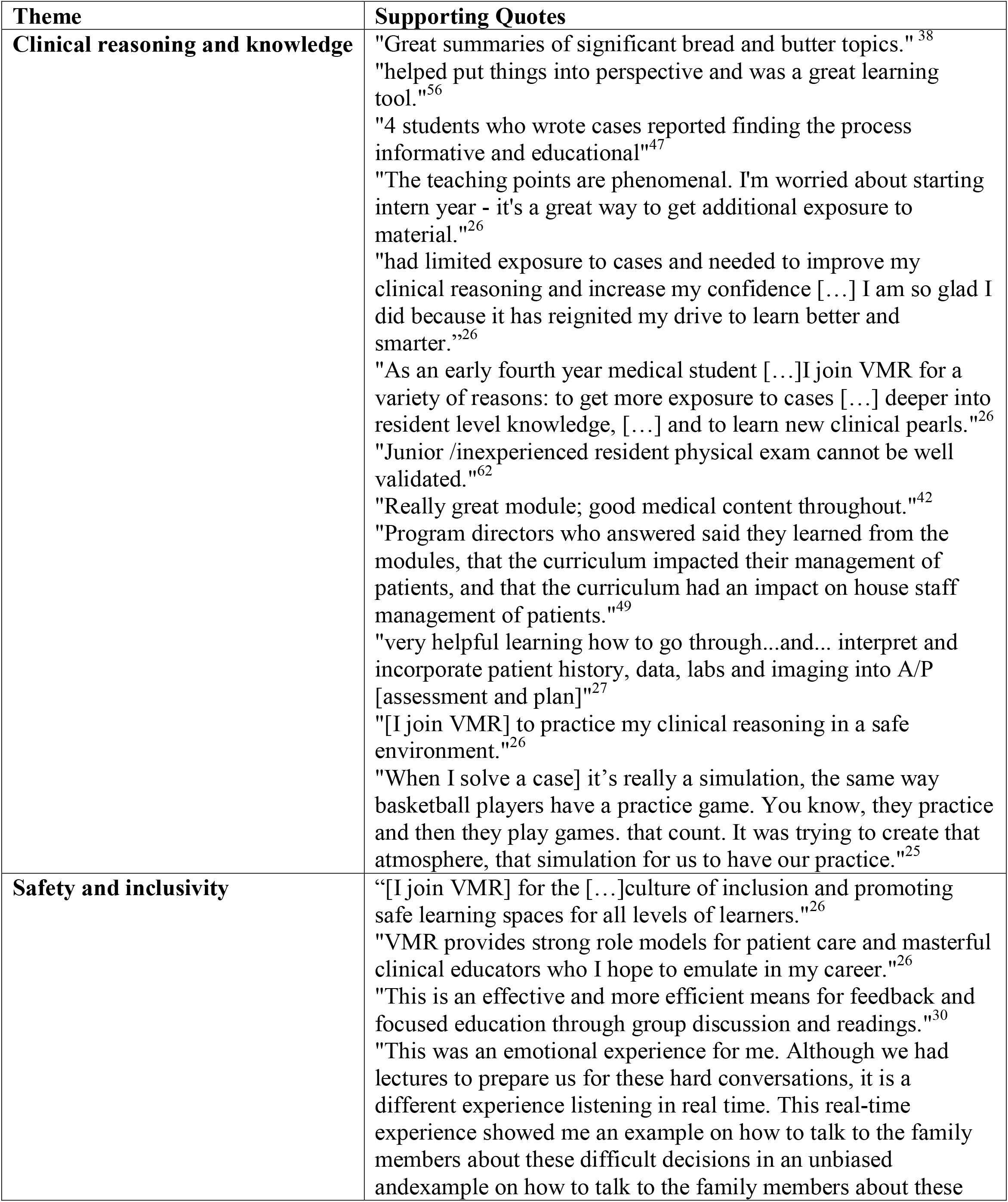
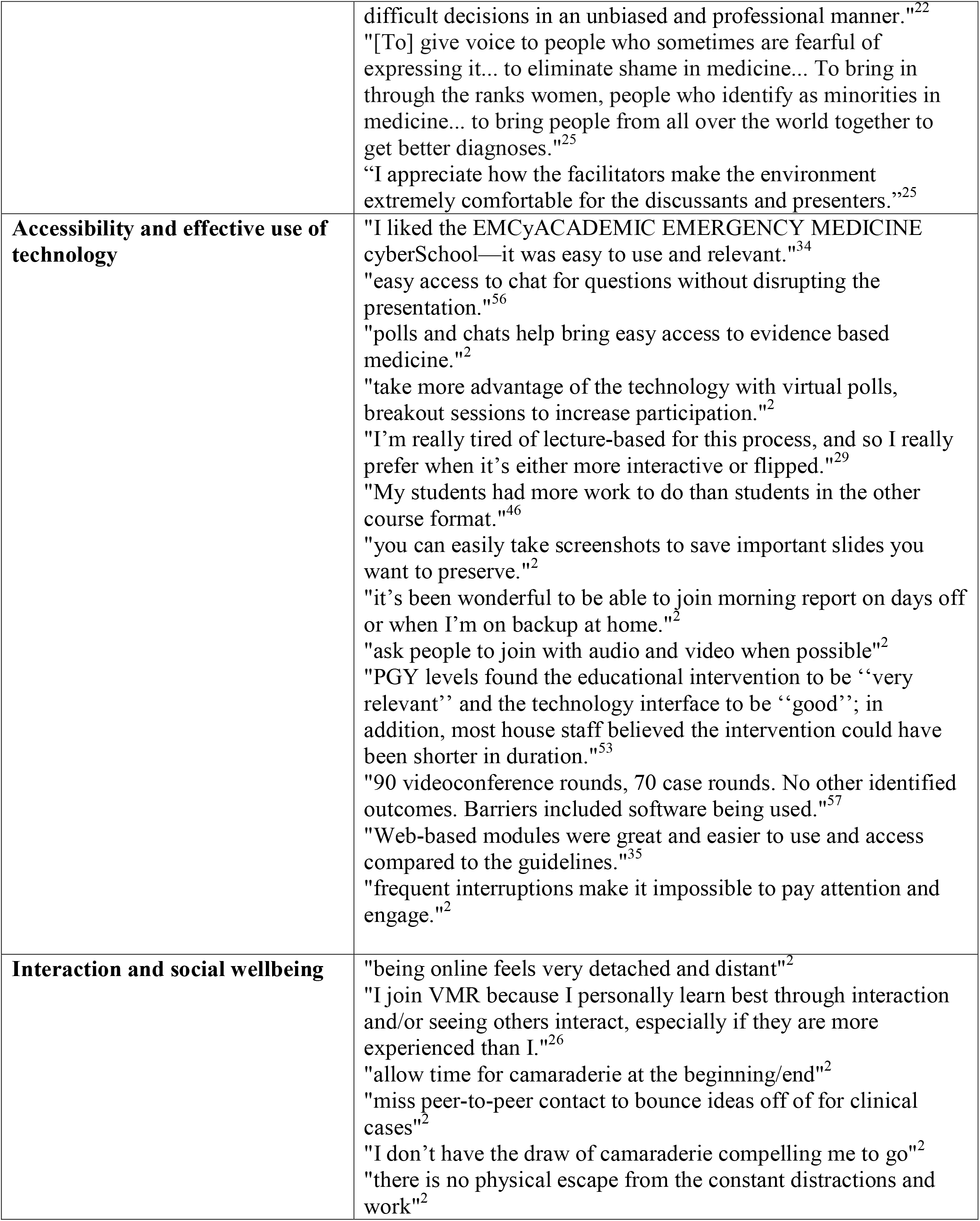
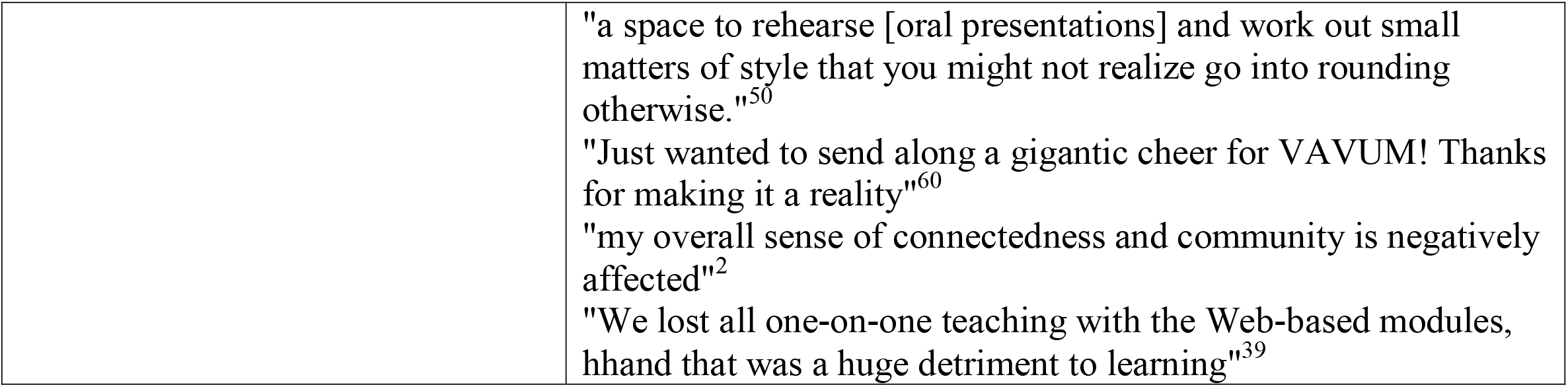
Thematic analysis.

#### Clinical reasoning and knowledge (Theme 1)

Most learners found VMR and case-based virtual teaching educational and helpful in reinforcing medical knowledge.^26,38,47,56^ Learners found VMR increased confidence with clinical reasoning and acquired valuable clinical pearls.^25,26^ Many respondents also enjoyed a simulation experience for practice.^26^ Some program directors responded that the VMR curriculum often improved house staff management of patients. On the other hand, learners voiced concerns about lack of validation of physical exams for junior learners without access to bedside teaching.^62^

#### Safety and inclusivity (Theme 2)

Learners found that VMR democratized the learning process, particularly through use of videoconferencing and podcasts, and the virtual format was found to be more learner-centred.^22,25,26,30^ Learners from diverse backgrounds who may have had less opportunities to speak in traditional settings found a safe platform to engage during VMR.^25^ Virtual morning report with videoconferencing platforms such as Zoom and Microsoft Teams allowed for easier access to the chat function for questions by learners who may have hesitated in traditional classroom settings.^2,25,26^ The use of diverse groups of facilitators and lecturers in podcasts was also appreciated by many participants.^25^ A central theme of feeling safe resonated with many participants in the virtual setting, which resulted in improved knowledge consolidation.^25,26^

#### Accessibility with technology (Theme 3)

Improved ability to access educational sessions from home was highlighted as a key positive theme. ^2,34^ Increased accessibility included having online repositories and capturing online slides with screenshots for later review.^2^ The use of chat boxes was again noted to be a helpful technological feature. ^2,56^ Although most comments were positive, challenges around efficiency with cumbersome technology were also noted.^46,53^ Most residents preferred shorter virtual teaching sessions and some struggled with the added burden of online passwords and websites for access.^57^ Some instructors also noted additional work with creating modules.^46^ Finally, some learners commented that technological interruptions such as poor Internet quality may impact learning.^2,53,57^

#### Interaction and social wellbeing (Theme 4)

Peer engagement and an interactive learning environment were highlighted as key priorities for learners.^2,26^ The switch to VMR reduced the ability to socialize with peers and learn from group discussions.^2^ Learners noted that they felt psychologically detached from colleagues, peers, and attendings.^2,39^ Residents noted that one way to improve peer engagement virtually would be to allow time for camaraderie either at the beginning or end of VMR sessions.^2^ It was also important to use video and audio capabilities to simulate a real setting to increase feelings of connectedness. ^2,60^ Learners suggested using virtual polls and team sessions to increase participation. ^2,50^ In general, learners preferred interactive case discussion with videoconferencing over lecture-based modules.^26,29^ Finally, the overall impact on social wellbeing was negative if virtual learning did not allow from escape from constant work related distractions while at home. ^2^

## Discussion

Our review of the literature demonstrated a gap in research on best practices and effectiveness of VMR with wide heterogeneity for learning and clinical outcomes. We found that learners benefitted from VMR due to reinforcing clinical knowledge with the flexibility of access from home, a safe and inclusive learning environment, and improved decision-making capacity.

However, learners found that VMR also reduced access to bedside teaching, patient interaction, and peer engagement, which led to lower social connectedness. Overall, VMR continued to be an effective tool for engaging learners, particularly with use of audiovisual applications for conducting group discussions during the COVID-19 pandemic. Use of Zoom or Microsoft Teams applications allowed for faculty and student interaction. Use of chatboxes and polls increased peer engagement and was an effective way to ask questions.

The literature on VMR does not have standardized frequency, format, length of time, or expected outcomes. Although the traditional morning report was initially instituted to improve patient handover^66^ and monitor daily patient care, there is very limited understanding of the impact on clinical outcomes.^67^ Similarly, our review found a paucity of research on the effectiveness of virtual morning report on patient and clinical outcomes. The morning report has undoubtedly transitioned into a form of teaching method tailored to specific residency program needs with a lesser role in active patient care.^54^ The recent literature after the COVID-19 pandemic demonstrated a widening of this gap whereby patient care and online teaching was often divorced from each other. This gap in patient-centered and bedside learning was noted by some learners and faculty members as a barrier of VMR.

Overall, our review faces challenges of limited controlled studies, heterogeneity of participants and programs, and small sample sizes within programs. We were also not able to capture all grey literature although attempts were made using Google Scholar to capture abstracts and proceedings, and two of the included citations are abstracts from grey literature. There is also no clear standard definition or format for VMR, and we expanded our definition to incorporate studies on virtual rounds or virtual case-based teaching. Many program evaluations also lack comparisons to traditional forms of morning report, which limit our understanding of comparative effectiveness.

We identified key areas for future directions in research. We need further studies exploring use of online platforms that allow for patient confidentiality, adhere to privacy and ethics, while also allowing for increased engagement between learners, faculty, and patients. Artificial intelligence (AI) use in medical education is an emerging area and can be leveraged to optimize adaptive learning and case simulation for learners for improved patient management.^54,69,70^ Use of randomized trials in medical education is also emerging and ten studies in this review explored this methodology. The key to success in ongoing virtual settings will be focusing on interactive formats and engagement between learners and faculty as identified as a key priority in this scoping review. Finally, assessing long term impacts of VMR on patient and clinical outcomes is necessary with longitudinal studies.

## Conclusions

Virtual morning report offers learners a safe and inclusive learning environment with benefits of expanded clinical knowledge, improved accessibility from home, and the ability to revisit cases later for reinforcing topics at the expense of reduced social connectedness.

## Data Availability

All data produced in the present study are available upon reasonable request to the authors.

## Acknowledgements

The authors thank Meagan Stanley, academic librarian at Western University for her help in establishing the search strategy.

## Competing Interests

All authors declare that they have no competing interests or disclosures.

## Funding

S. Sarma received the Dr. Parveen Wasi Medical Education Research Award from McMaster University, of which this scoping review is a component.

## Ethical approval

Reported as not applicable.

## Notes

### Competing Interest Statement

The authors have declared no competing interest.

### Funding Statement

This study did not receive any funding.

