## Supplemental Appendix for "A scoping review of virtual morning report and outcomes in Canada and the United States"

**Supplementary Appendix 1**

**Search Strategy: EMBASE and OVIDMedline**

--------------------------------------------------------------------------------

1	computer-assisted instruction.tw. or Computer-Assisted Instruction/ (12288)

2	(online learning or distance education).tw. or Education,Distance/ (7185)

3	online education.tw. (774)

4	web-based learning.tw. (401)

5	case-based learning.tw. (575)

6	learning module*.tw. (1447)

7	morning report*.tw. or Morning Report/ (1447)

8	virtual learning.tw. (525)

9	medical residents*.tw. or "Internship and Residency"/ (54592)

10	medical internship*.tw. (109)

11	medical education.tw. or Education,Medical/ (88600)

12	1 or 2 or 3 or 4 or 5 or 6 or 7 or 8 (22120)

13	9 or 10 or 11 (135075)

14	12 and 13 (3937)

15	internal medicine.tw. (25712)

16	14 and 15 (174)

17	limit 16 to english language (164)

***************************

Additional search keywords for Google Scholar and PubMed:

“virtual morning report”

“virtual case-based teaching”

“online morning report”

“online case-based teaching”
